# Antibiotic Stewardship in Lower Urinary Tract Infections among the Elderly at a District General Hospital

**DOI:** 10.1101/2025.08.29.25334484

**Authors:** Priyankaran Mithrakumar, Milton Barua

## Abstract

**Objectives:** This study aimed to evaluate the management of lower urinary tract infections (LUTIs) in elderly inpatients, to determine the prevalence of inappropriate treatment of asymptomatic bacteriuria (ASB), and to assess the impact of targeted educational interventions on antibiotic prescribing accuracy.

**Settings:** An acute general hospital in East Midlands of the United Kingdom.

**Participants:** Seventy-seven patients aged 65 and older who have had a urine culture due to suspected diagnosis of lower urinary tract infection.

**Design:** Prospective case series of emergency hospital admissions collected over an 8-month period to acute elderly medicine wards. Implementation of two educational interventions at the second and third month of the study period.

**Methods:** Data relating to demographics, catheterization status, presenting symptoms, urine dipstick usage, culture collection methods, and antibiotic prescription patterns were obtained from electronic and paper records as part of audit number 1688 registered at the hospital trust. Interventions included the display of an informational poster based on NICE guidelines and a dedicated educational teaching session for clinicians. In total, 19 patients were involved pre-intervention. A total of 58 patients were involved post-intervention. Among the post intervention cohort, 32 patients were involved post-poster intervention, and 17 patients were involved post-teaching intervention. Several months following intervention, 9 patients were involved in the study.

**Results:** Initial findings revealed suboptimal adherence to national guidelines, with 50% (n=5) of asymptomatic patients with negative cultures inappropriately prescribed antibiotics. Following the poster intervention, prescribing was found to be three and a half times more appropriate for LUTIs than the pre-intervention period (OR=3.61, 95%CI 1.08 to 12.03, (⍰^2^ =4.56, p=0.033). Compared to the pre-intervention period antibiotic prescribing was found to be five times (OR=5.05, 95%CI 0.96 to 26.66, ⍰^2^=3.91, p=0.048) more appropriate after all educational interventions implemented. Catheter-associated infections also showed a significant reduction, albeit with poor statistical correlation.

**Conclusion:** Targeted educational interventions, including informational posters and teaching sessions, were observed to improve appropriate diagnosis and antibiotic prescribing for LUTIs and ASB in elderly hospitalized patients within the scope of this study.

## Introduction

Lower urinary tract infections (LUTIs) represent a prevalent category of infections in both community and healthcare settings. Accurate diagnosis of a LUTI requires a combination of relevant clinical signs and symptoms, ideally supported by growth of bacteria in urine cultures. Commonly, elderly patients with no symptoms can also have growth of bacteria in their urine cultures and this is called asymptomatic bacteriuria (ASB). There is strong evidence to support that fact that treatment of ASB with antibiotic use does not improve outcome in long-term care settings except in pregnancy and patients undergoing invasive urological procedures as explained in Petty LA., 2019 [1]. Despite national guidelines recommending against antibiotic therapy in ASB, use remains persistently high in long-term care facilities such as hospital and residential homes as described by Trevino SE et al., 2016 [2].

The primary objectives of this study were to quantify the current LUTI diagnostic pathway, antibiotic prescribing pattern, implement and study the effectiveness of simple interventions to improve clinical effectiveness in LUTI management in a single hospital setting. Indeed, studies such as Lee et al., 2018 [3] have demonstrated the effectiveness of educational intervention in decreasing antibiotic treatment in long-term care residents. The novelty of this study is in the analysis and implementation of such intervention in an acute general hospital in the east midlands of the United Kingdom. This study also explores the effectiveness of a teaching session on prescribing outcomes which goes further to the poster and pocket card-based strategy implemented by Lee et al., 2018 [3] in Saskatchewan, Canada. This study also contributed to quality improvement by ensuring that LUTI investigation and management in medical wards adhered to established guidelines as published in NICE guideline NG112 [4].

The incidence of healthcare-associated infections, such as *Clostridioides difficile* (*C. difficile*), has been rising nationally and locally in England. Woodford HJ et al., 2009 [5] found that among 265 patients in an acute general hospital in the northwest of England, 43.4% of patients were over diagnosed with LUTI. The treatment given to these patients varied and led to *C. difficile* infections in 8% of cases, falls in 4% and methicillin-resistant staphylococcus aureus (MSRA) infection in 3%. Prior observations showed a disproportionately high incidence of antibiotic-associated infections, specifically *C. difficile*, among elderly medical patients, alongside instances of inappropriate antibiotic prescribing at our local institution based in the east midlands of the United Kingdom. A particular area of clinical uncertainty for healthcare providers has been antibiotic prescribing in cases of bacteriuria identified on urine culture and the accurate diagnosis of LUTIs in delirious patients as described in Woodford HJ et al., 2009 [5]. By improving LUTI diagnostic pathways and treatment strategies we hoped to reduce healthcare associated infection, in the acute general hospital we were based.

## Methods

This study was conducted in several general medicine wards and included data from 77 patients who had a documented suspected lower urinary tract infection (LUTI) and had undergone urinary cultures due to the same. Key data points collected comprised the ward audited, patient demographics (age, gender, and ethnicity), catheterisation status, presenting symptoms; whether a urine dipstick was performed, the method of culture collection and details of antibiotics initiated following the culture report. Additionally, details regarding the specific antibiotics prescribed were recorded. Medical staff working on the ward, were encouraged to contribute to data collection towards this project as an incentive to improve their knowledge as demonstrated in Gimbel et al., 2017 [6].

As a quality improvement initiative, this audit was registered with the hospital’s governance framework and was deemed to fall under standard clinical practice evaluation, thereby not requiring formal review by an Institutional Review Board or Research Ethics Committee. All patient data collected were prospectively and retrospectively extracted from routine clinical records and were anonymized or de-identified prior to analysis to reduce confirmation bias.

The Inclusion criteria for the study were patients aged 65 years or older admitted to the local hospital with a suspected lower urinary tract infection and had urinary cultures done. It was standard practice in our institution that all patients with a suspected lower urinary tract infection had urinary cultures done. Exclusion criteria included patients less than 65 years old, pregnant patients, LUTIs with sepsis, and LUTIs complicated by other infective conditions requiring antibiotics. Data collection from medical wards was facilitated via a standardised multiple-choice proforma, shown in appendix 1, which was delivered electronically to data collectors via their smart phones. What constitutes positive lower urinary tract symptoms and positive urinary culture, as defined by NICE Summary on LUTIs in over 65, NICE, n.d. [7], were explicitly stated for data collectors on the proforma to ensure consistent and accurate information retrieval during data collection as demonstrated in figure 1. In addition, data collection was delegated to medical students and qualified doctors working in the wards under investigation to reduce selection bias. When data collectors had doubts about the data collected, the principal author was available to provide clarification about LUTI diagnostic criteria or signpost to further educational resources.

**Figure 1:**
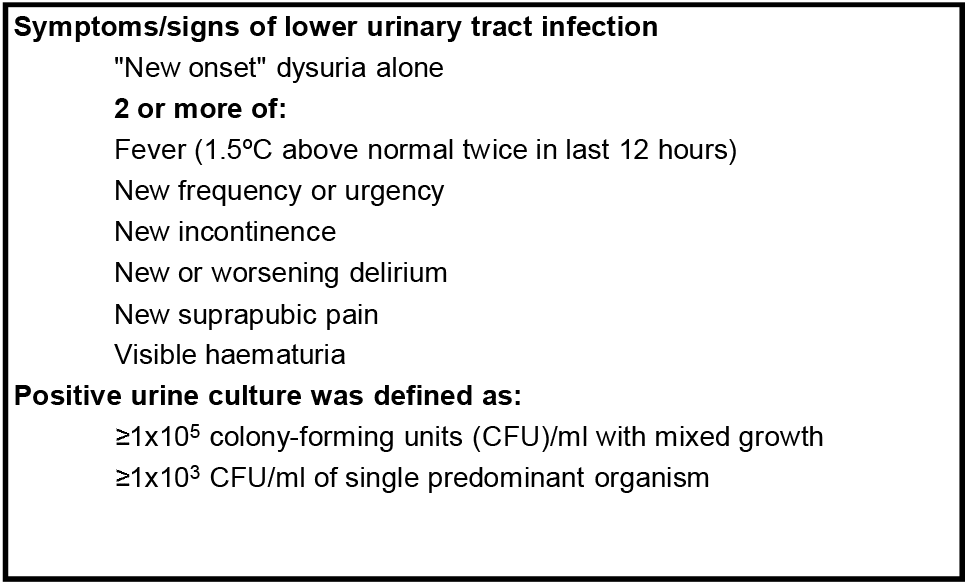
NICE diagnostic criteria for Urinary Tract Infection in those 16 and over in the United Kingdom as published in NICE Summary on LUTIs in over 65, NICE, n.d. [7]

Figure 2 shows a brief methodological flow diagram of the timeline for data collection and implementation of interventions. During the pre-intervention period, data was collected on 19 patients admitted to the elderly general medicine wards at the institution. We ensured that patients were studied from a mix of different medical wards to ensure that local factors to a specific ward did not impact the generalisability of our data. Baseline findings revealed that antibiotic prescribing practices at the local hospital did not consistently align with national standards as published by NICE guideline NG112 [4]. Our primary objective was to identify local prescribing gaps and implement targeted stewardship interventions. Despite expanding data collection to different wards and involving multiple data collectors the study samples were small and limited to specific wards, due to constraints caused by real-world ward admissions, availability of patient cohort during the study period and other logistical constraints. A preliminary intervention needed to be implemented despite limited data due to significant findings in the shortfall of patient care which constituted a patient safety concern. In addition, due to the patient safety concerns, a concurrent control group was not feasible as interventions were implemented ward-wide to mitigate risks associated with shortfalls in LUTI management. Following discussion with the lead consultant, posters derived from the diagnostic decision tool for adults over 65 years with suspected uncomplicated LUTI, published in UK Health Security Agency & NHS England., 2025 [8] were placed in key clinical decision-making areas such as the doctors’ office, ward round workstations and nursing stations. Appendix 2 shows the design and layout of the poster that was implemented.

**Figure 2:**
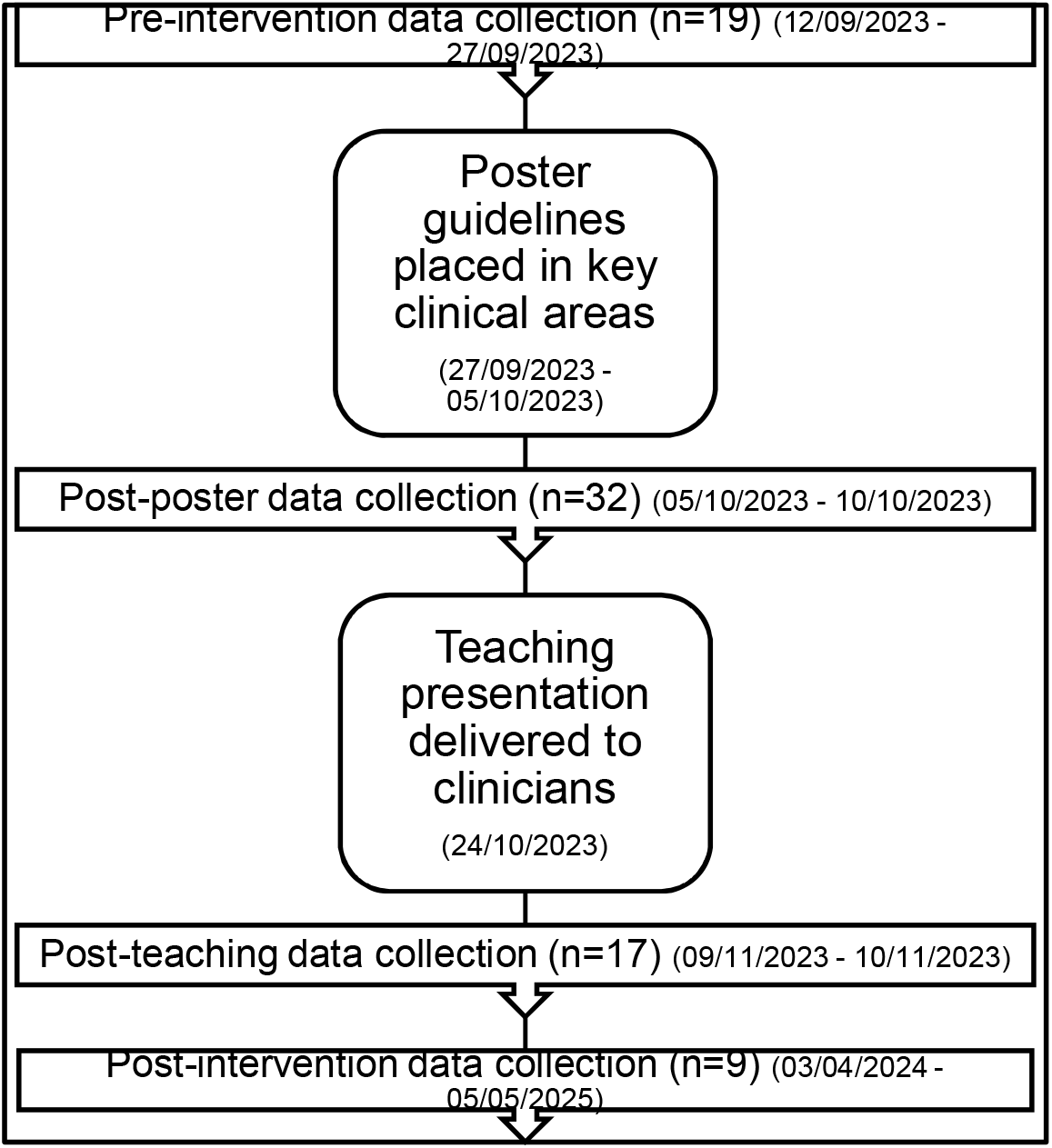
Methodology flow diagram summarising the intervention timeline and sample sizes across study phases

A further cycle of data was collected from 32 patients in October 2024 to measure the impact of the intervention of posters on the management of LUTIs. We did not intend the study samples between intervention to be statistically equivalent but as sequential snapshots showing temporal impact of contemporaneous intervention lead by the second primary author, a consultant in elderly medicine and implemented by the corresponding author and the ward team. The study findings were initially intended to be exploratory, highlighting the temporal association between quality improvement initiatives. On 24th October 2023, a teaching session was held for all resident doctors, senior doctors and medical staff working in the department of elderly medicine. This teaching session was one of four, delivered to staff as part of a teaching programme developed to improve care to patients over the age of 65 years. The teaching was focused on three main areas: diagnostic criteria for LUTIs, antibiotic prescribing guidelines and principles in urinary catheter care in elderly patients (age > 65), as published by NICE Summary on LUTIs in over 65, NICE, n.d. [7]. The teaching session ran for approximately 30 minutes in the department of elderly medicine and included interactive quizzes for the audience. Slides of the teaching session on LUTIs, relevant to this study, are shown in appendix 3 and were delivered by the corresponding author who is a fully qualified doctor under the supervision of the second primary author, a consultant in elderly care medicine. Feedback was obtained from a validated questionnaire designed by the medical education department of the institution shown in appendix 4.

A further cycle of data was collected from 17 patients in the elderly medicine wards immediately following the teaching session and are shown in the results section. The findings of these multiple audits were presented to the infection prevention and control (IPC) team based in the institution on November 3^rd^, 2023. The infection prevention team regarded the presentation as highly beneficial but recommended enhancements to strengthen clinician benchmarking and enable the collection of more precise, clinically relevant data for future quality improvement initiatives.

### Data analysis

The primary outcome of this study was to evaluate the quality of LUTI management in elderly medicine wards following implementation of targeted educational interventions. From the data collected appropriate antibiotic prescribing was classified as asymptomatic patients who were either culture negative without antibiotics or culture positive without antibiotics to demonstrate appropriate omission of antibiotics in asymptomatic bacteriuria (ASB). In symptomatic patient’s appropriate antibiotic prescribing was classified as patients who were culture negative without antibiotics, representing cases whose symptoms did not directly correlate with active LUTI, and those who were culture positive on antibiotics getting appropriate treatment for LUTI.

Inappropriate antibiotic prescribing was classified as asymptomatic patients who were culture negative but on antibiotics and those who are culture positive and on antibiotics representing inappropriate treatment of ASB. Symptomatic patients with inappropriate antibiotic prescribing were classified as those who were culture negative and on antibiotics representing the cohort of patients on antibiotic therapy who have symptoms associated with other frailty syndromes but not to active LUTI. Symptomatic patients who were culture positive without antibiotics were also classified into the inappropriate treatment category because they did not receive clinically indicated treatment.

The temporal association pre- and post-intervention, such as poster and teaching, on appropriate antibiotic prescribing was studied by collecting data pre and post intervention. The odds of appropriate antibiotic prescribing pre and post intervention was compared with odds of inappropriate prescribing during the same period to obtain an odds ratio for each intervention of improving antibiotic prescribing. As antibiotic prescribing is a categorical variable Chi-Square (⍰^2^) tests were used to study whether observed differences between pre and post intervention were statistically significant. Logistic regression was used to evaluate whether potential confounders such as age, gender, ethnicity, catheter use, and ward had significant influence on appropriate antibiotic prescribing.

The secondary outcome of this study was to evaluate whether educational intervention had any impact on ward-based catheter management practices. Chi-square (⍰^2^) tests were used to evaluate whether ward based educational interventions had any significant statistical impact on good catheter management practice.

## Results

### Pre-intervention study

Figure 3 presents the distribution of urinary culture outcomes among elderly patients’ pre-intervention, stratified by symptomatic status.

**Figure 3:**
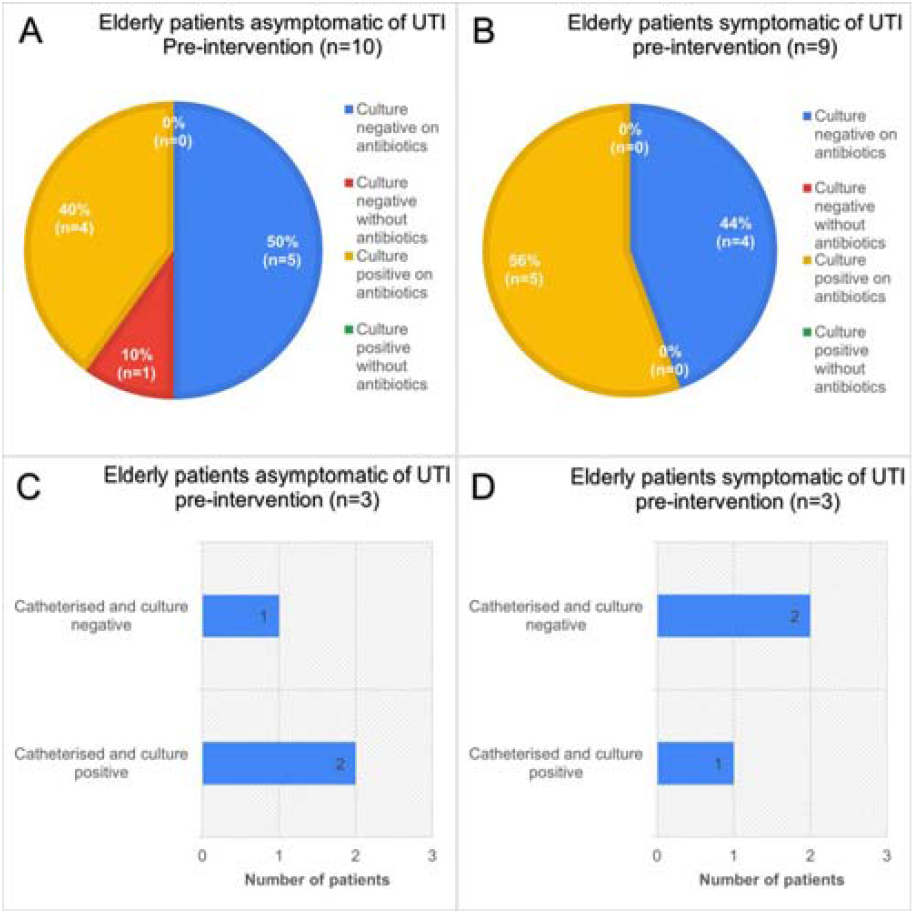
Data obtained from a pre-intervention audit of antibiotic prescribing practice, in a total of 19 patients aged > 65 years, admitted to elderly medicine wards. <u>A: The number of patients asymptomatic of LUTI, B: Patient symptomatic of LUTI, C/D: catheterisation status in 6 patients</u>.

Among, asymptomatic patients (n=10) (Figure 3A), half (50%, n=5) had a negative culture while receiving antibiotics. A further 40% (n=4) were culture positive while on antibiotics. Only one patient (10%) had a negative culture without antibiotics, and none were culture positive without antibiotic use.

In contrast, among symptomatic patients (n=9) (Figure 3B), the majority (56%, n=5) demonstrated culture positivity while receiving antibiotics. The remaining 44% (n=4) were culture negative on antibiotics. No symptomatic patients were found to be culture negative without antibiotics or culture positive without antibiotics.

Figures 3C and 3D illustrate catheterisation status in a subset of asymptomatic (n=3) and symptomatic (n=3) patients. Of the asymptomatic group, two patients were catheterised with a culture-positive result, while one was catheterised with a culture-negative result. Within the symptomatic cohort two patients were catheterised and culture negative, while one was catheterised and culture positive, suggesting urinary symptoms likely due to catheter irritation.

Overall, these findings suggest that a substantial proportion of both symptomatic and asymptomatic elderly patients were receiving antibiotics despite negative culture results, while positive cultures were observed predominantly in patients on antibiotics. These findings were likely due to empiric antibiotic initiation prior to availability of culture results, erroneous cultures, or treatment directed toward urinary symptoms not attributable to LUTI as described by Rousham et al., 2019 [9]. Catheterisation was associated with both culture-positive and culture-negative findings across groups, likely reflecting asymptomatic bacteriuria (ASB) due to suboptimal catheter practices.

### Post-poster intervention

Following the poster intervention, culture–treatment patterns in older adults showed clearer clinical separation by symptom status as shown in figure 4.

**Figure 4:**
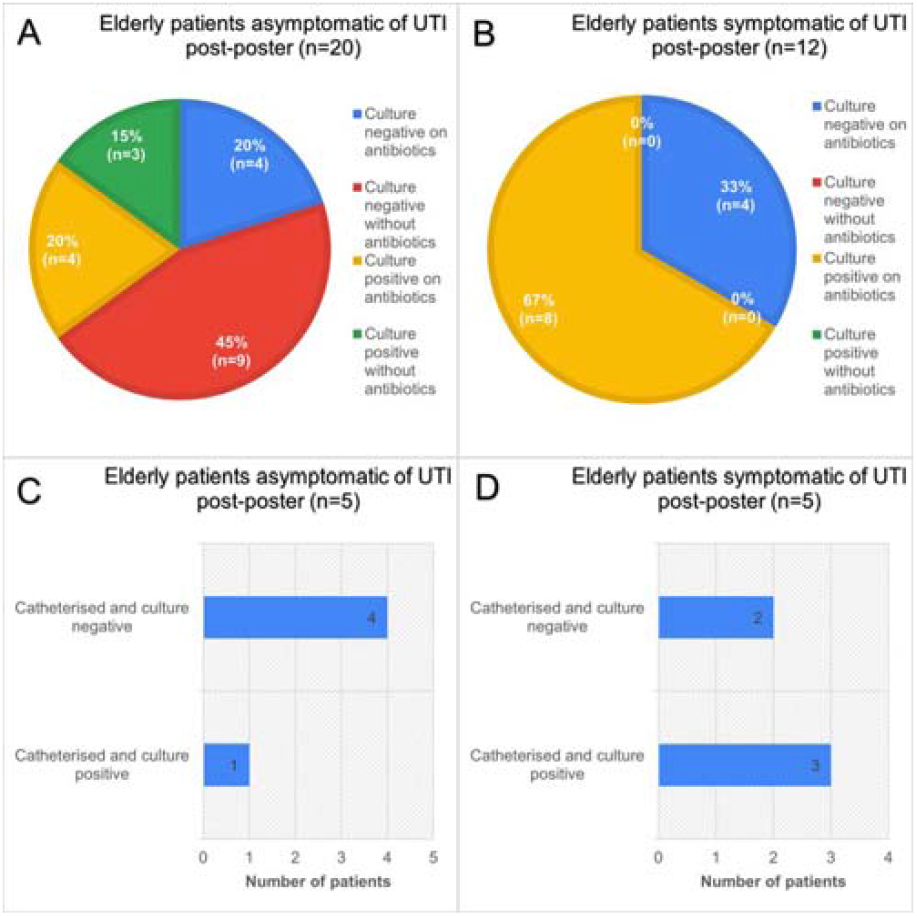
Data obtained during post poster-intervention period during September 2023, of a total of 32 patients aged > 65 years, admitted to elderly medicine wards. <u>A: The number of patients asymptomatic of LUTI, B: Patient symptomatic of LUTI, C/D: catheterisation status in 10 patients</u>.

Among asymptomatic patients (n=20; Fig. 4A), nearly half were culture-negative and not on antibiotics (45%, n=9). A further one fifth were culture-negative while receiving antibiotics (20%, n=4). Culture-positive results were less frequent and split between those on antibiotics (20%, n=4) and not on antibiotics (15%, n=3). Clinically, most asymptomatic individuals were either not treated or had antibiotics withdrawn in the context of negative cultures, with a smaller group continuing therapy despite no microbiological evidence. In symptomatic patients (n=12; Fig. 4B), two thirds were culture-positive on antibiotics (67%, n=8), while one third were culture-negative on antibiotics (33%, n=4).

Catheterised subgroups reflected the diagnostic challenges of bacteriuria. Among asymptomatic catheterised patients (n=5; Fig. 4C), four were culture-negative and one culture-positive, aligning with colonisation being common yet often non-pathogenic. Among symptomatic catheterised patients (n=5; Fig. 4D), three were culture-positive and two culture-negative, indicating that symptoms alone did not reliably distinguish infection from catheter-related irritation or other causes.

Overall, the post-poster data depict more better antibiotic stewardship. Antibiotic prescribing following poster intervention was found to be three and a half times more appropriate for LUTIs than the pre-intervention period (OR=3.61, 95%CI 1.08 to 12.03) and this difference was found to be statistically significant (⍰^2^ =4.56, p=0.033). Despite this finding, there was no statistical improvement in catheter management practice following poster intervention (⍰^2^ =0.311, p=0.55). These findings support the view that antibiotic administration was appropriately guided by the poster’s reinforced definition of LUTI as described in Funaro JR, et al., 2021 [10] but did not address poor catheter care.

### Post-teaching intervention

After the presentation, management patterns showed a clearer alignment with symptoms and culture results as shown in figure 5.

**Figure 5:**
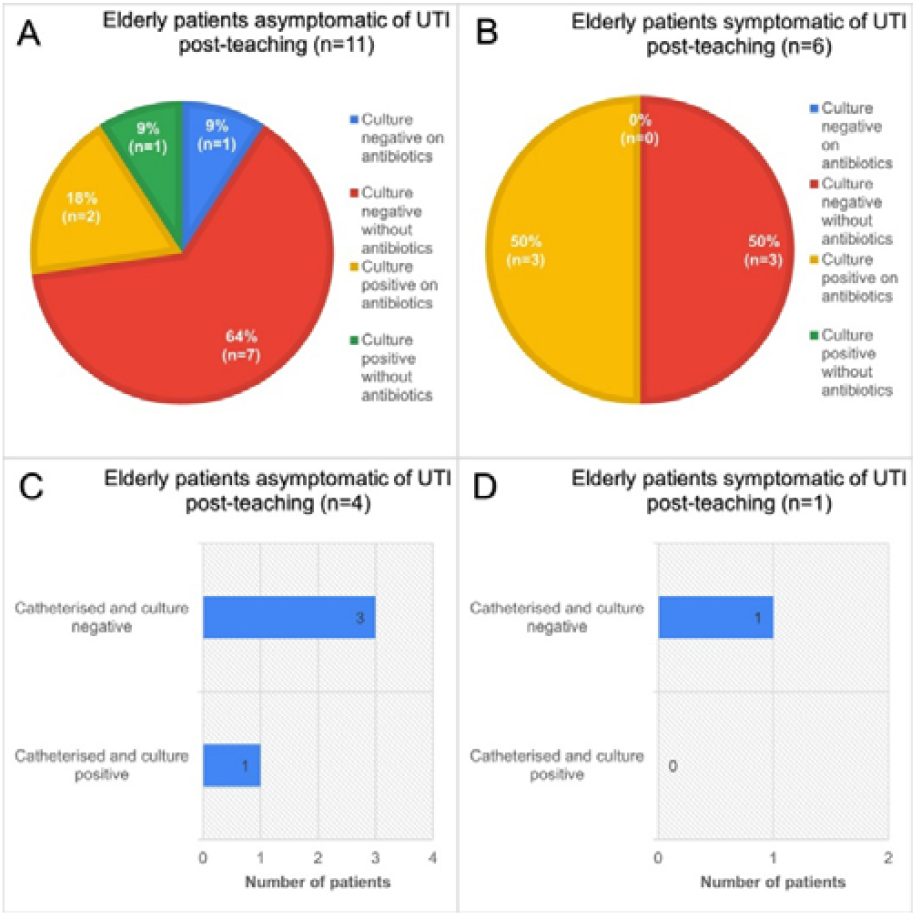
Data obtained during post teaching-intervention period during October 202,3 of a total of 17 patients aged > 65 years, admitted to elderly medicine wards. <u>A: The number of patients asymptomatic of LUTI, B: Patient symptomatic of LUTI, C/D: catheterisation status in 5 patients</u>.

Among asymptomatic older adults (n=11; Fig. 5A), most had negative cultures without antibiotics (64%, n=7), indicating restraint from treatment in the absence of microbiological evidence. Only one patient (9%) was culture-negative while on antibiotics. Culture positivity was uncommon and split between those on antibiotics (18%, n=2) and not on antibiotics (9%, n=1). In symptomatic patients (n=6; Fig. 5B), half were culture-positive on antibiotics (50%, n=3), consistent with timely therapy for confirmed infection and the other half were culture-negative without antibiotics, indicating that a proportion of symptomatic presentations were not supported by culture and were not treated empirically, likely reflecting cases where symptoms such as acute confusion or altered mental status were deemed unrelated to LUTI as explained in Saukko PM et al., 2019 [11]. Statistical analysis showed no significant increase (⍰^2^ =2.06, p=0.151) but did show that appropriate prescribing was three times more likely (OR=2.8, 95%CI 0.66 to 11.79) after the teaching session than after posters placed in key clinical areas.

Among asymptomatic catheterised patients (n=4; Fig. 5C), three were culture-negative and one culture-positive, reflecting frequent colonisation without infection and supporting non-treatment in the absence of symptoms. The single symptomatic catheterised patient (n=1; Fig. 5D) had a negative culture, underscoring that catheter-related symptoms may have non-infective causes. There was no statistically significant difference (⍰^2^=1.53, p=0.22) in catheter management practice following the teaching session, likely due to insufficient sample size.

Six participants evaluated the LUTI-management teaching session as shown in figure 6. In terms of perceived quality (Fig. 6A), most respondents rated the session highly: 50% (n=3) outstanding and 33% (n=2) above average; the remainder rated it average (17%, n=1). No ratings were below average or poor. In terms of confidence to apply learning (Fig. 6B), after the session, 83% (n=5) reported feeling very confident applying LUTI-management knowledge and 17% (n=1) felt extremely confident. No participant reported low or no confidence. In terms of self-rated knowledge (Fig. 6C), before teaching, 3/7 participants assessed their knowledge as average, 2/7 as above average and 1/7 reported excellent knowledge. After teaching, 1/7 rated their knowledge average, 2/7 above average and 2/7 excellent. Overall, 57% moved to above average or higher post-teaching, indicating a clear perceived learning gain. In terms of qualitative feedback (Fig. 6D), free-text comments described a good presentation on a useful topic and relevant to elderly care. Participants suggested offering the session to a broader audience to maximise impact.

**Figure 6:**
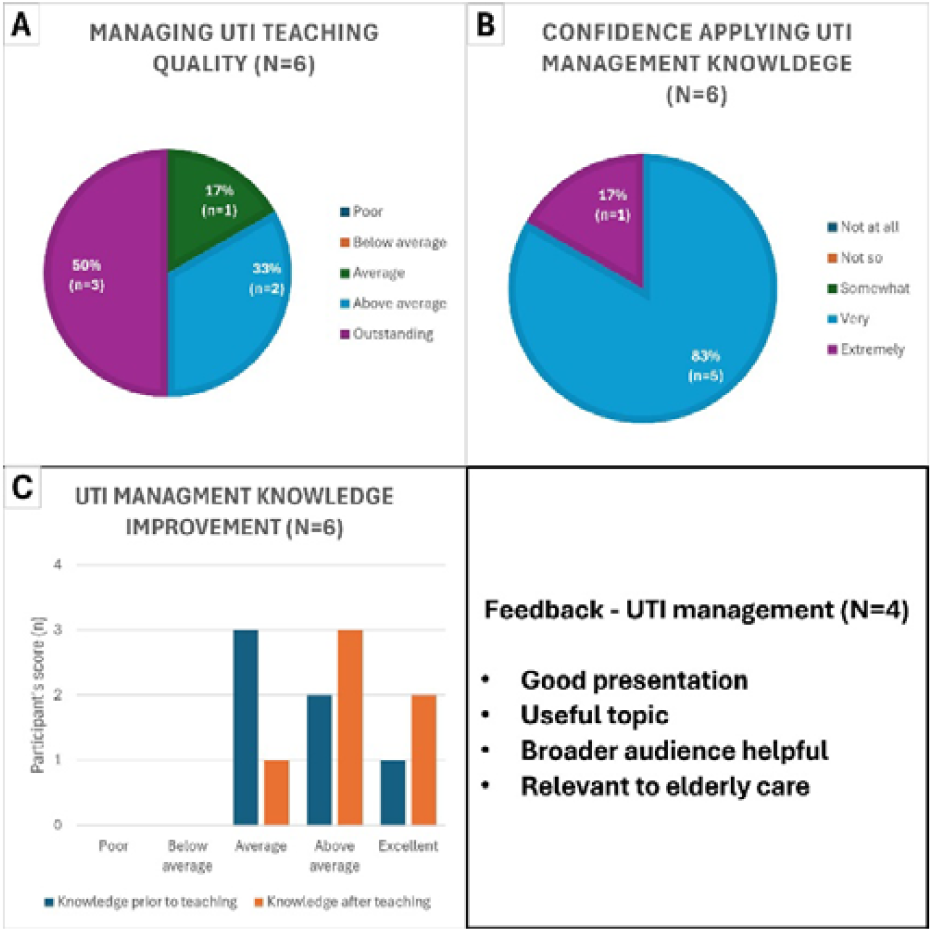
Responses, from a total of 6 participants to all questions in the feedback questionnaire for LUTI management teaching session. <u>A/B/C: Quantitative feedback. D: Qualitative feedback from 4 participants who responded</u>.

In summary, attendees judged the teaching to be high quality, reported high confidence in applying the content, and demonstrated an upward shift in self-assessed knowledge, supporting the educational effectiveness and practical relevance of the LUTI-management session. Overall, the post-teaching data depict greater antibiotic stewardship in asymptomatic elders with negative cultures, continued appropriate treatment of culture-confirmed symptomatic infections, and persistent diagnostic ambiguity in catheterised patients.

### Post-intervention study

Post-intervention, culture–treatment patterns in older adults showed greater separation by symptom status, albeit in small numbers as shown in figure 7.

**Figure 7:**
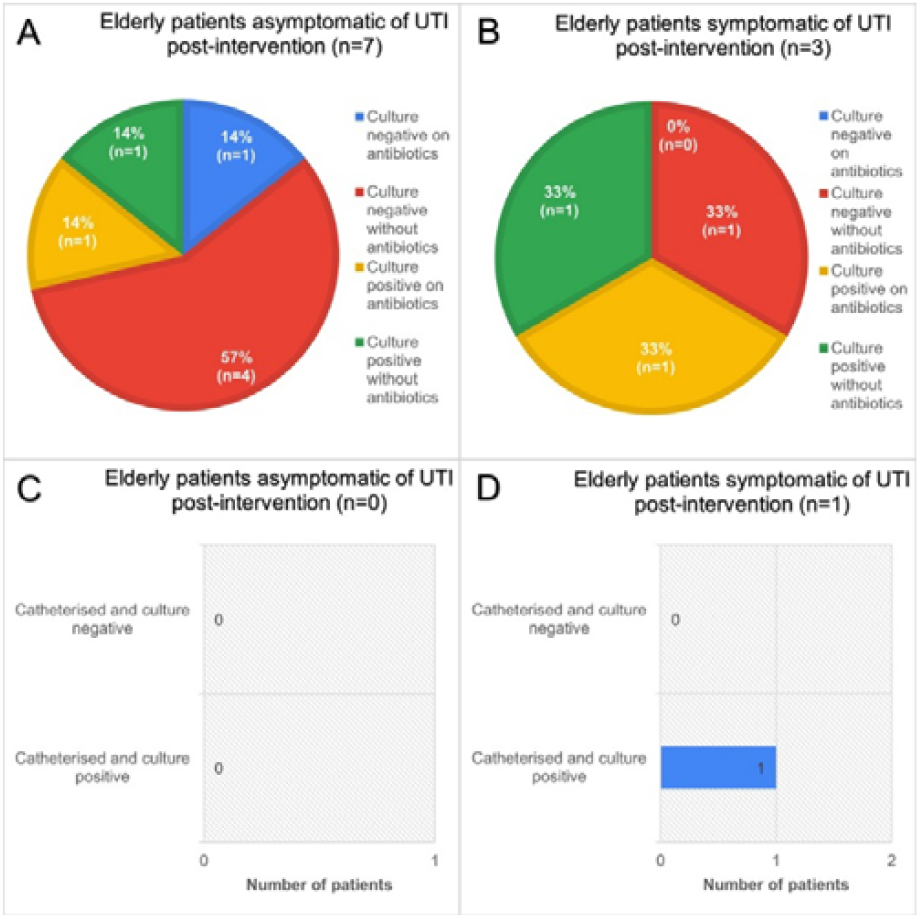
Data obtained during post-intervention period in April 2024 and beyond, of a total of 9 patients aged > 65 years, admitted to elderly medicine wards. <u>A: The number of patients asymptomatic of LUTI, B: Patient symptomatic of LUTI, C/D: catheterisation status of 1 patient</u>.

Among asymptomatic patients (n=7; Fig. 7A), most had negative cultures without antibiotics (57%, n=4), indicating restraint from treatment when infection was not supported microbiologically. The remaining cases were evenly split: culture-negative on antibiotics (14%, n=1), culture-positive on antibiotics (14%, n=1), and culture-positive without antibiotics (14%, n=1). Thus, unnecessary antibiotic exposure among asymptomatic individuals was limited to a single culture-negative case, while bacteriuria—when detected—did not uniformly prompt treatment.

In symptomatic patients (n=2; Fig. 7B), one patient was culture positive on antibiotics, another culture positive without antibiotics and another culture negative without antibiotics. Overall, two thirds of symptomatic post intervention patients were treated appropriately. Catheterised subgroups were minimal; there were no asymptomatic catheterised patients during this period (Fig. 7C). The single symptomatic catheterised patient (n=1; Fig. 7D) was culture-positive, aligning with catheter-associated infection as a potential driver of symptoms.

Overall, the post-intervention data limit inferences owing to very small symptomatic samples and absence of asymptomatic catheterised cases. Despite this finding compared to the pre-intervention period antibiotic prescribing was found to be five times (OR=5.05, 95%CI 0.96 to 26.66) more appropriate in the post-intervention period several months after teaching sessions. The improvement in antibiotic stewardship from the pre-to post-intervention period was found to be statistically significant (⍰^2^ =3.91, p=0.048), suggesting that posters and the teaching session had a positive meaningful impact on the diagnosis and management of LUTI in patients aged 65 or over.

Due to the temporal nature if this case series it was essential to rule out the impact of confounding factors on the outcomes of the study. Additional data was collected alongside this study of the presumed confounding factors such as age, gender, ethnicity, catheter-use and ward location. A logistic regression analysis was used to check whether there were any statistically significant correlations between appropriate antibiotic prescribing and any of the measured confounding variables. Figure 8 shows a forest plot of the odd ratios of confounding variables and their significance on appropriate prescribing of antibiotics to elderly inpatients.

**Figure 8:**
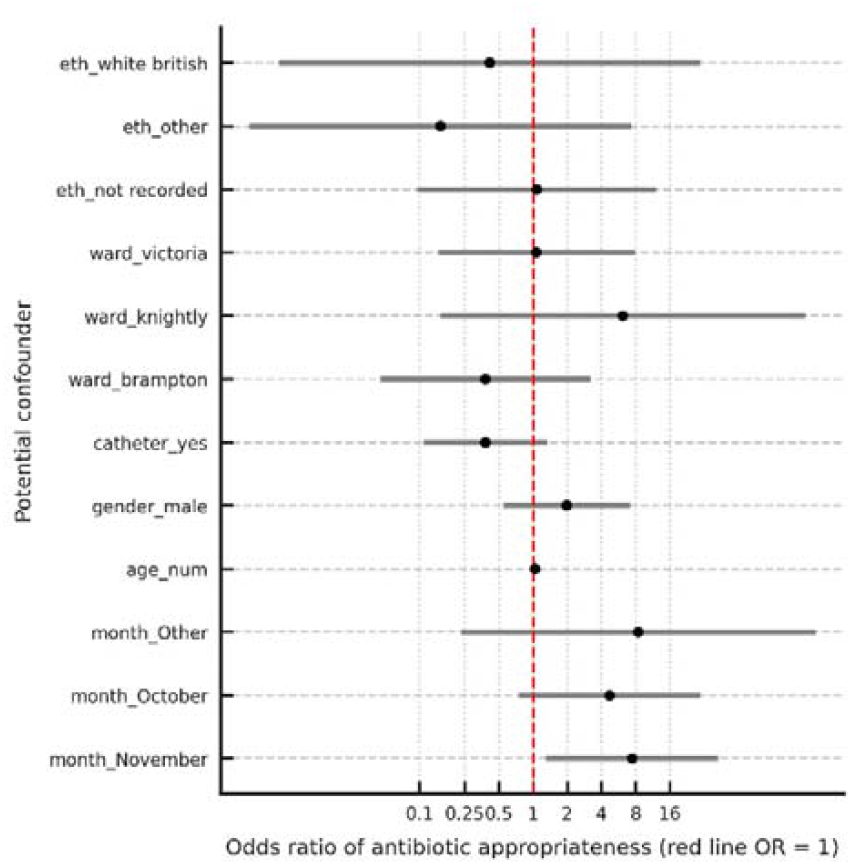
Forest plot of odds ratios and confidence intervals (CI) obtained through Logistic regression to study temporal trends in appropriate antibiotic prescribing for UTI in adult 65 and older; to adjust for potential confounders (age, gender, ward, catheter status and ethnicity)

Figure 8 shows that all presumed confounders did not have statistically significant effect on the odds of appropriate antibiotic prescribing during the period of study. Additionally, it is likely that the short, defined timeframes of each data collection cycle, as shown in figure 2, may have reduced the influence of seasonal infection patterns and staff turnover on prescribing practices and infection rates. As shown in figure 8 a statistically significant correlation was noticed only between the month of data collection and odds of appropriate antibiotic prescribing with November showing a significant improvement in practice compared to the baseline pre-intervention month of September.

## Discussion

This quality improvement project aimed to assess and improve antibiotic prescribing practices for lower urinary tract infections (LUTIs) and asymptomatic bacteriuria (ASB) in elderly patients admitted to general internal medicine wards at a district general hospital. The findings demonstrate an observable positive impact of targeted educational interventions on clinician behaviour, leading to more appropriate antibiotic use.

### Study strengths

The baseline data revealed multiple areas where local prescribing practices deviated from national standards, particularly regarding the inappropriate treatment of ASB. A substantial proportion of asymptomatic elderly patients were inappropriately prescribed antibiotics, often for presumed LUTIs when symptoms were more likely attributable to frailty syndromes such as delirium. This reflects wider concerns in antimicrobial stewardship, where misdiagnosis and unnecessary antibiotic prescribing contribute to antimicrobial resistance and adverse outcomes, including *C. difficile* infection as explained in Rousham et al., 2019 [9]. The persistently high incidence of *C. difficile* nationally and locally underscores the urgent need for interventions to reduce inappropriate prescribing as explained in Echaiz JF et al., 2015 [12].

The interventions implemented: an informational poster and a dedicated teaching session, contributed to improving clinicians’ understanding and adherence to appropriate diagnostic and prescribing criteria. Immediate (OR=3.61, 95%CI 1.08 to 12.03, ⍰^2^=4.56, p=0.033) and sustained improvements (OR=5.05, 95%CI 0.96 to 26.66, ⍰^2^ =3.91, p=0.048) were observed in the ability of clinicians to distinguish LUTI symptoms from other manifestations of frailty and administer appropriate antibiotic treatments. Qualitative feedback from teaching sessions further supported their utility in clarifying complex diagnostic scenarios, especially in cases involving ASB and catheter-associated infections as demonstrated in Lee et al., 2018 [3].

The study also provided important insights into the management of catheterised patients. The finding that some asymptomatic catheterised patients had positive urine cultures reflects the challenge of distinguishing colonisation from true infection in this group as explained in El Moussaoui., 2025 [13]. Early results suggested suboptimal catheter practices, but subsequent reductions in asymptomatic positive cultures and catheter-associated urinary tract infections (CAUTIs) indicated an improvement in clinical practice albeit poor statistical power (⍰^2^ =1.53, p=0.22). However, an initial rise in symptomatic catheterised patients with positive urine cultures suggested that catheter care protocols were not consistently applied, likely because the initial poster intervention did not explicitly advise on catheter changes during suspected CAUTIs. The observed correlation between LUTI symptoms and bacteriuria in catheterised patients also underscores the central role of clinical assessment alongside laboratory results in this complex patient population as explained in Shimoni Z et al., 2020 [14].

### Study weaknesses

This project was conducted at a single site, which may reduce generalisability to other healthcare settings. The relatively small sample size of 77 patients, although adequate for audit purposes, may compromise the statistical power and generalisability of findings to other health care settings as demonstrated by Faber and Fonseca., 2014 [15]. Due to the urgency of needing to address shortfalls in antibiotic prescribing to ensure patient safety, poster and teaching interventions had to be implemented at fixed time periods regardless of sufficient patient recruitment during the pre and post intervention periods to address clinical significance or data reliability. Data collection relied on standardized proformas, which in turn depended on the accuracy of clinical documentation as explained in Echaiz JF et al., 2015 [12], but partially addressed bias caused by inter-rater reliability in data interpretation during collection. Furthermore, while the study strongly suggests that improved prescribing practices would reduce *C. difficile* infections and other adverse events, such outcomes were not directly measured. Longer-term follow-up would be beneficial to confirm the sustainability of improvements and to assess downstream effects on patient safety and antimicrobial resistance patterns. Another key limitation of this study was the significant imbalance across study phases which raises concerns about comparative analysis validity. This uneven distribution makes it difficult to draw meaningful conclusions about intervention effectiveness and may introduce selection bias. The imbalance across study phases was due to the fixed timelines during which the poster and teaching were implemented. Limited number of patients meeting the inclusion criteria and availability of data collectors during the specified phases of the study lead to varying number of patients recruited into the study during the different phases of the study.

Although this study shows promising preliminary observations, the pilot nature of this work, warrants further investigation in larger, controlled studies. Small subgroups of patients in the post intervention group may lead to overinterpretation of findings that could be due to chance alone. The small sample sizes in the post intervention group also make it difficult to apply statistical testing. Due to competing commitments of the authors and data collectors in this study, this was a prospective observational study performed using a contemporaneous sampling method. Arguably the lack of a control group, fails to address the potential confounding variables such as seasonal infection patterns, concurrent quality improvement initiatives, staff turnover, or changes in hospital policies that may influence antibiotic prescribing practices. The authors attempted to control for some of these variables by registering the audit with the clinical governance department ensuring that this work was unique during the period of study; implemented a standardised proforma to ensure that staff turnover did not affect the quality of data collection and ensured that hospital policies did not change during the study period by liaising with the infection prevention and control (IPC) team. The authors used chi-squared statistical tests and logistic regression analyses to reduce bias caused by comparative analysis validity and confounding factors, however it is advisable that future projects should explicitly measure and adjust for such confounding variables.

Despite these limitations, several key recommendations emerge from the findings from this study conducted in a single institution. Clinicians would benefit from further training to improve recognition of LUTI symptomatology, particularly in elderly or delirious patients with impaired communication. Ward teams should be educated about the high prevalence of ASB in catheterised patients and reminded to prioritise catheter removal or replacement rather than antibiotic prescribing when clinically appropriate. Diagnostic accuracy could also be improved by incorporating blood inflammatory markers and urine microscopy into the assessment of elderly patients with communication difficulties as explained in El Moussaoui., 2025 [13]. In addition, broad-spectrum antibiotics should be reviewed and de-escalated within 24–48 hours of initiation, ideally through collaboration with pharmacy teams, to counteract unfavourable prescribing practices, particularly those originating in emergency departments as written in Shrestha J et., 2023 [16]. This study provides real-world clinical relevance, multi-modal interventions and shows promising results that are easily reproducible within the ward environment. The conduct of this study contributed to the improvement of care for high-risk elderly populations and informs future studies focusing on this pertinent clinical need.

## Conclusion

This study provided preliminary evidence of improved prescribing trends temporally associated with educational interventions. The findings are hypothesis-generating and warrant further investigation in the form of larger multicentre studies to confirm generalisability. While the study does not have the statistical power of a multicentre trial, our primary aim was to identify local prescribing gaps and implement targeted stewardship interventions. The implementation of an informational poster and a dedicated teaching session led to a notable improvement in clinician behaviour, resulting in more judicious antibiotic prescribing practices. This improved understanding directly contributed to a reduction in the inappropriate treatment of asymptomatic bacteriuria, a key component of effective antibiotic stewardship. Ultimately, these efforts foster a healthcare environment that prioritises patient safety by minimising unnecessary antibiotic exposure, thereby mitigating the risks of antibiotic resistance and adverse drug reactions such as *C. difficile* infection.

## Data Availability

All data produced in the present study are available upon reasonable request to the authors

## Appendix

**Appendix 1:**
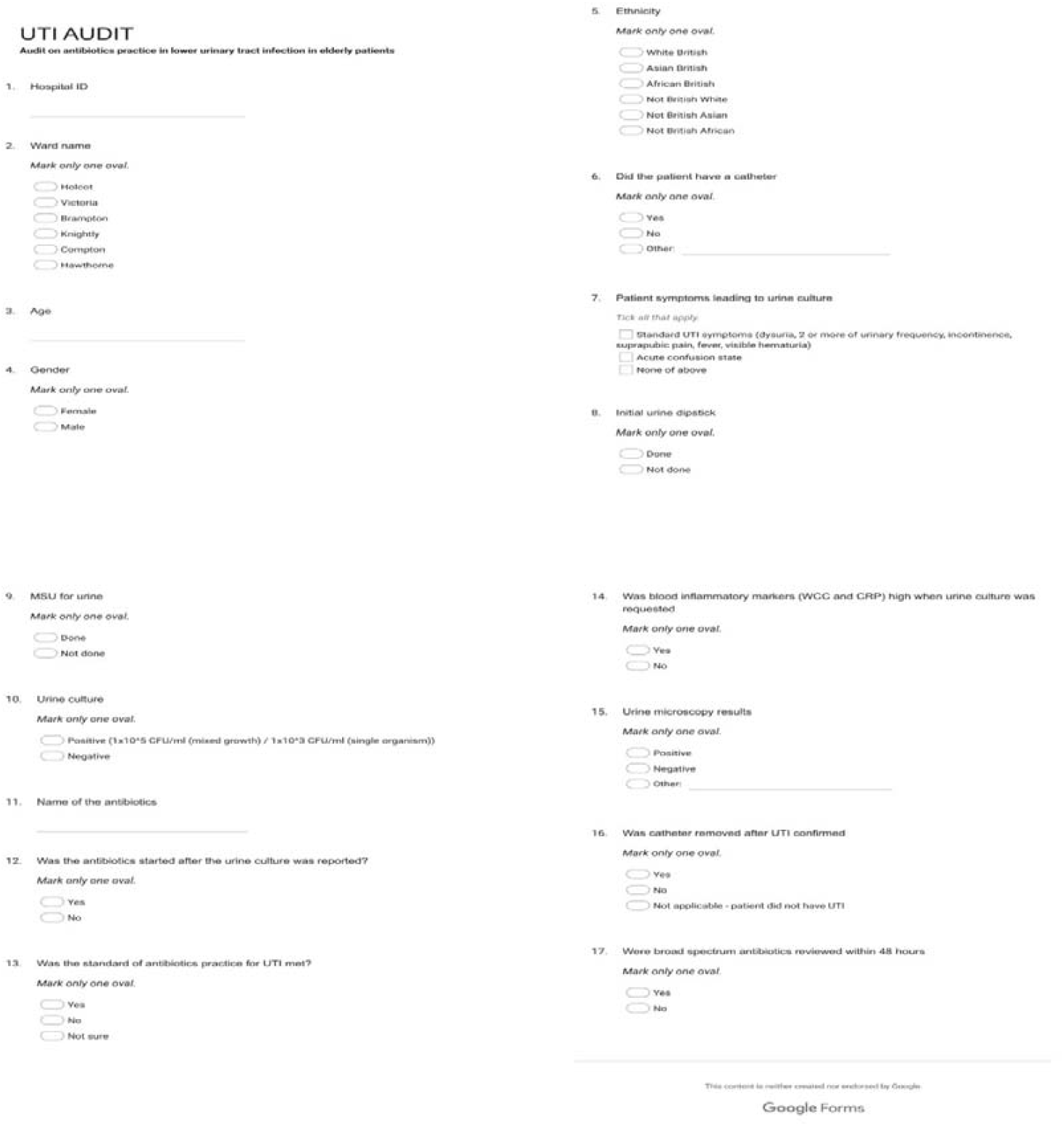
The electronic proforma that was used to collect data on UTI management trends at the health institution

**Appendix 2:**
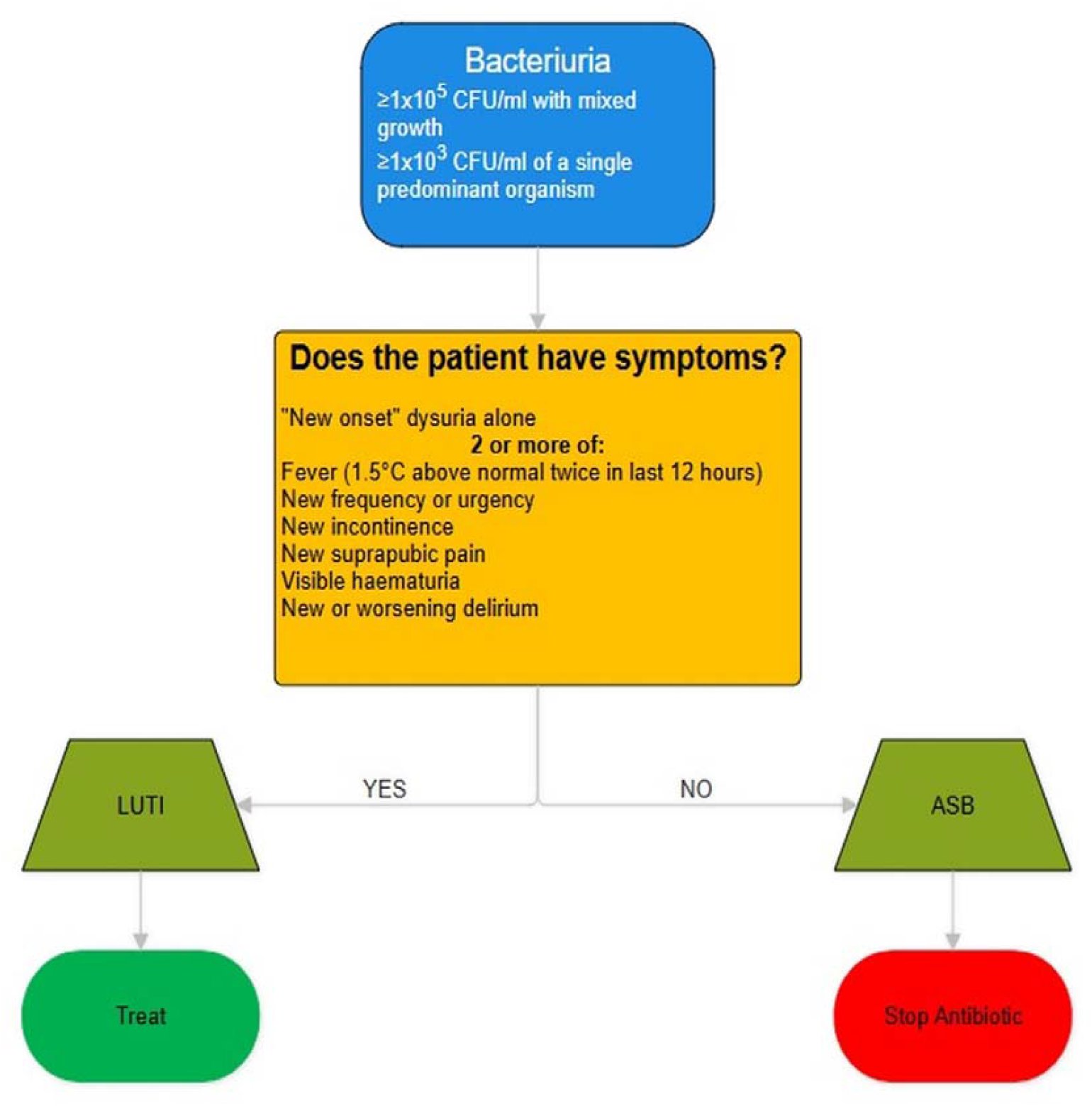
Design and layout of the poster that was implemented as part of the post-intervention in this study adapted from UK Health Security Agency & NHS England., 2025 [8]

**Appendix 3:**
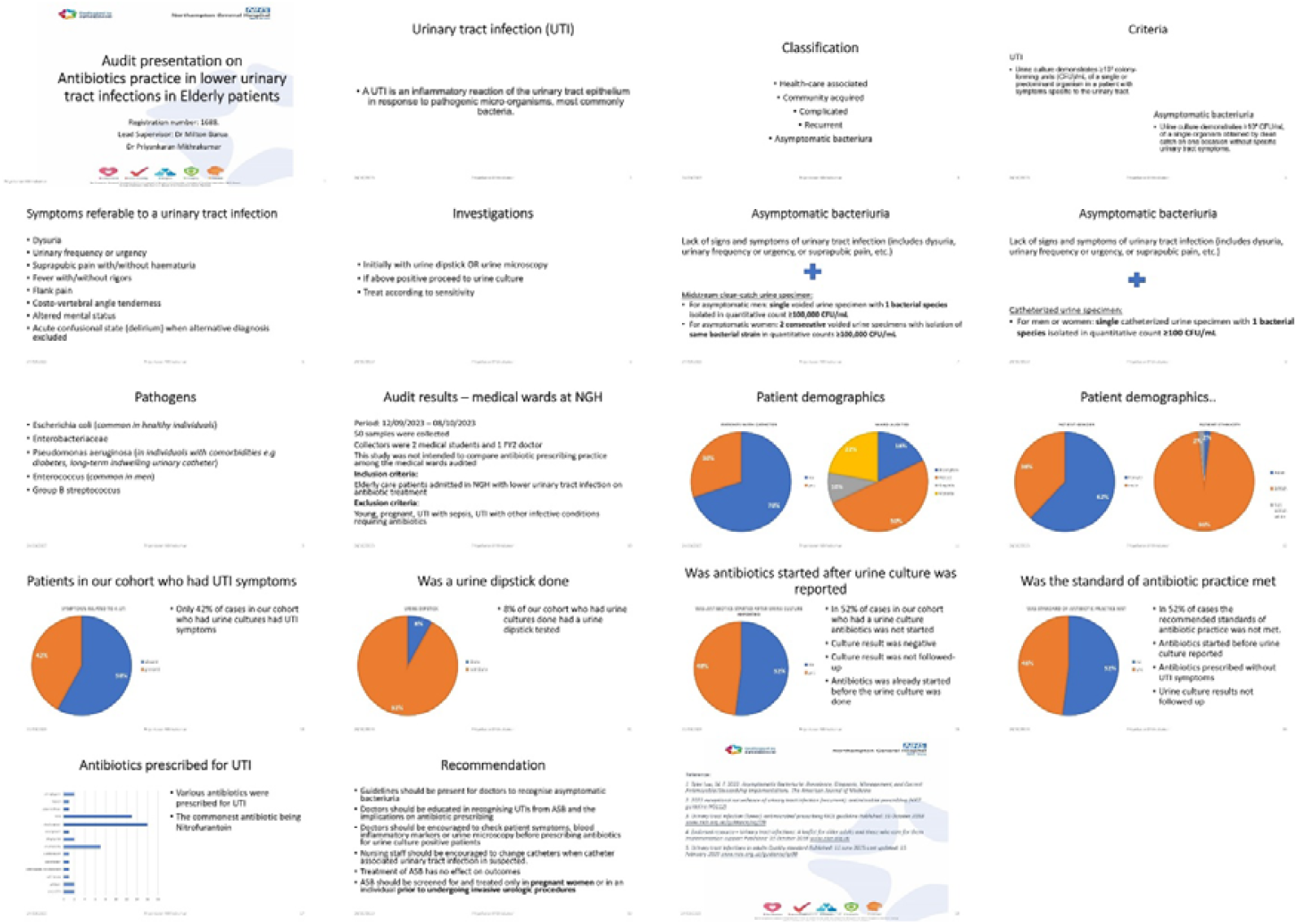
Slides of the teaching presentation that was used as part of the post-intervention in this study.

**Appendix 4:**
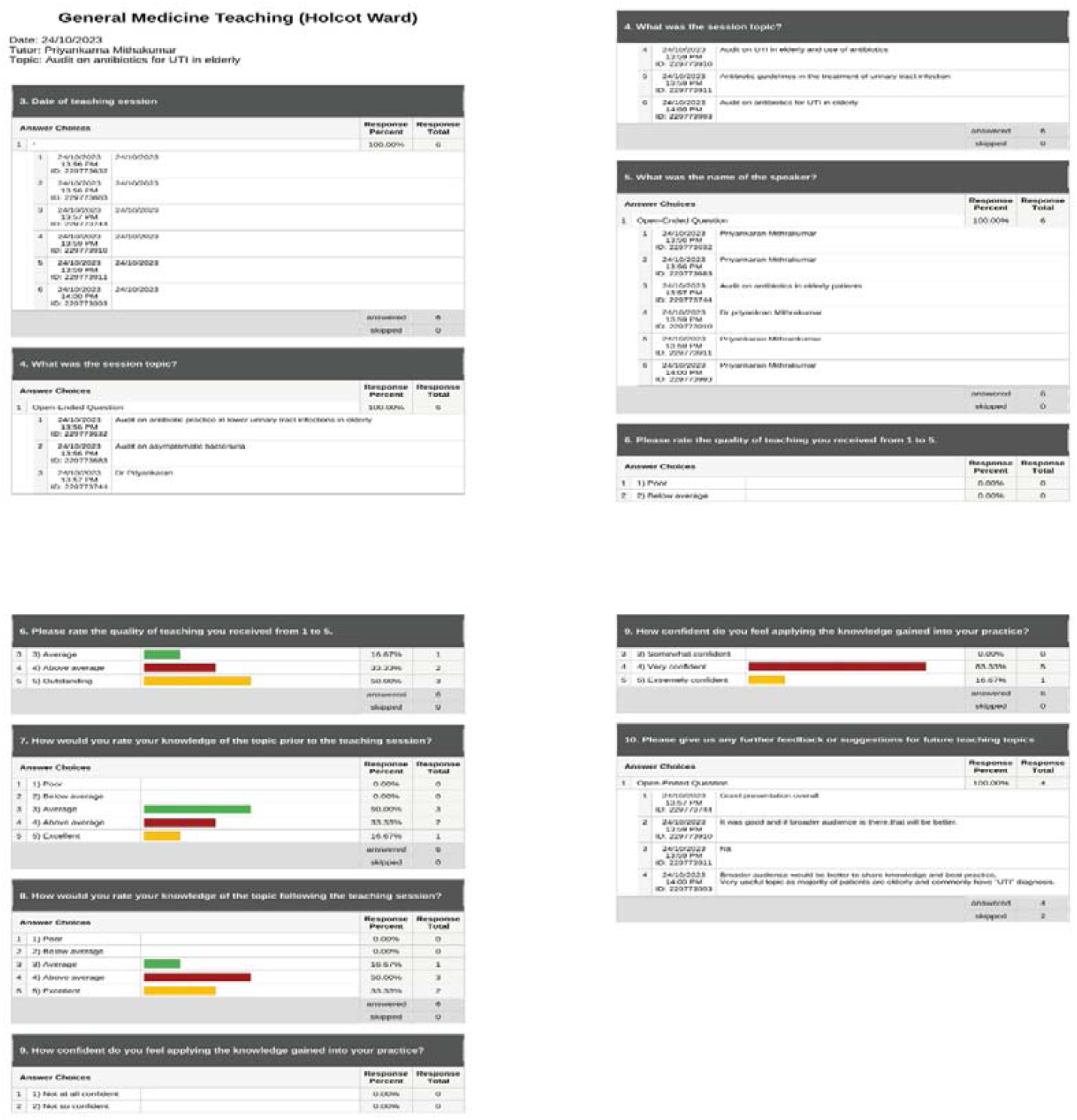
Complete feedback questionnaire for UTI management teaching session carried out on 24th October 2023 in the elderly medicine department of the institution being studied.

